# Prospective Audit of Hand-Scrubbing Practices in an Orthopedic Operating Theatre in a Regional Headquarter Hospital of Northern Pakistan

**DOI:** 10.64898/2026.03.14.26348091

**Authors:** Sohail Rehman, Zakria Rathore, Mehdi Ali Mehdivi, Nasir Hussain, Laiba Uroosh

**Author notes:** **Corresponding Author:** Sohail Rehman.

## Abstract

**Background:** Hand hygiene remains a cornerstone of infection prevention in surgical practice, particularly in orthopedic operating theatres where inadequate aseptic technique can increase the risk of surgical site infections and implant-related complications. Despite well-established recommendations from the World Health Organization (WHO) regarding proper surgical hand-scrubbing techniques, compliance in many healthcare settings remains inconsistent. Clinical audits provide a structured approach to evaluating adherence to such guidelines and implementing targeted improvements. This study aimed to assess baseline hand-scrubbing practices in an orthopedic operating theatre at a regional hospital in northern Pakistan and evaluate the impact of educational interventions on compliance with WHO standards.

**Methods:** A prospective closed-loop clinical audit was conducted in the orthopedic operating theatre of Regional Headquarter Hospital Skardu, Pakistan, from December 1, 2025, to February 1, 2026. Approximately 40 healthcare personnel, including consultants, residents, nurses, and operating theatre assistants, participated in the audit. Baseline hand-scrubbing practices were observed during routine surgical sessions using a structured checklist based on WHO hand hygiene guidelines. Following the baseline assessment, educational interventions were introduced, including live demonstrations of correct hand-scrubbing techniques and placement of visual reminder posters in the scrub area. Post-intervention compliance was re-evaluated using the same checklist. Compliance rates before and after the intervention were compared using appropriate statistical analysis, with significance set at p < 0.05.

**Results:** Baseline observations revealed suboptimal compliance with recommended hand-scrubbing standards, particularly with regard to scrubbing duration, coverage of all hand surfaces, and proper drying technique. Following the educational intervention, significant improvements were observed across all evaluated components. Compliance with scrubbing duration of at least two minutes increased from 45% to 90%, coverage of all hand surfaces improved from 50% to 88%, proper antiseptic usage increased from 60% to 93%, and correct drying technique improved from 55% to 87%. Adherence to overall aseptic protocol also increased from 70% to 95%. All observed improvements were statistically significant (p < 0.001).

**Conclusions:** This prospective clinical audit demonstrates that structured educational interventions, including live demonstrations and visual reminders, can significantly improve compliance with recommended hand-scrubbing practices in orthopedic operating theatres. Regular audits combined with targeted educational strategies represent practical and cost-effective measures for improving infection control practices and enhancing patient safety in surgical settings.

## Introduction

Hand hygiene is universally recognized as the cornerstone of infection prevention in healthcare settings, particularly in surgical environments [1]. In orthopedic surgery, inadequate hand scrubbing is associated with increased risk of surgical site infections, prosthetic joint contamination, and postoperative morbidity [2,3]. Despite clear guidelines from the World Health Organization (WHO) and other bodies outlining recommended hand-scrubbing techniques-including duration, coverage, and antiseptic use-compliance in many operating theatres remains inconsistent, especially in resource-limited settings [4,5].

Clinical audits provide a structured method to assess adherence to these guidelines and implement interventions to improve practice. Prospective, closed-loop audits are particularly valuable as they allow for baseline assessment, targeted interventions, and re-evaluation to measure improvement [6].

This audit aimed to evaluate the baseline hand-scrubbing practices of staff in an orthopedic operating theatre at the Regional Headquarter Hospital in Skardu, Pakistan, implement educational interventions including live demonstrations and visual reminders, and assess the subsequent improvement in compliance with WHO standards.

## Materials & Methods

### Study Design and Setting

A prospective, closed-loop clinical audit was conducted from December 1, 2025, to February 1, 2026, in the orthopedic operating theatre of the Regional Headquarter Hospital, Skardu, Pakistan. The audit included a multidisciplinary team of approximately 40 participants, comprising consultants, residents, nurses, and operating theatre assistants (OTAs).

### Audit Standards

The WHO hand-scrubbing technique was used as the benchmark standard [1]. Key components included:

Adequate duration (≥2-3 minutes)

Complete coverage of all hand and forearm surfaces

Proper use of antiseptic solutions and drying methods

Adherence to aseptic protocol during donning of gloves

### Baseline Assessment

Hand-scrubbing practices were observed during routine surgical sessions. Observers used a structured checklist to record adherence to the standard protocol. Hand-scrubbing practices were observed during routine surgical sessions using a structured observation checklist derived from the World Health Organization hand hygiene guidelines. The complete observation pro forma used for data collection is presented in Appendix Table *2*.Participants were unaware of specific audit criteria to minimize bias.

### Educational Intervention

Following baseline assessment, a multifaceted educational intervention was implemented:

Live demonstration sessions led by senior surgical staff, illustrating correct hand-scrubbing technique

Posters and visual cues placed strategically in the scrub area to reinforce technique

Interactive discussion addressing common errors and emphasizing infection control benefits

### Re-assessment

After the intervention, hand-scrubbing practices were re-evaluated using the same observation checklist. Improvements in compliance were quantified as percentages of adherence for each component.

### Statistical Analysis

Descriptive statistics were calculated, including proportions of compliance for each hand-scrubbing element. Differences between pre- and post-intervention adherence were assessed using the Chi-square test or Fisher’s exact test where appropriate. Statistical significance was set at p < 0.05. Data were analyzed using SPSS version 26.

## Results

At baseline, overall compliance with WHO hand-scrubbing standards was suboptimal, particularly for duration, coverage of all hand surfaces, and proper drying technique. Consultants demonstrated higher adherence than residents and support staff, although deviations were still observed across all categories.

Following the educational intervention, marked improvements were observed in nearly all components of hand hygiene. Compliance with scrubbing duration, thoroughness of coverage, and correct antiseptic usage increased significantly (p < 0.01). Early initiation of post-scrub glove donning and maintenance of aseptic technique were also enhanced.

**Table 1:** summarizes pre- and post-intervention compliance for key hand-scrubbing elements.

| Hand-Scrubbing Component | Baseline Compliance (%) | Post-Intervention Compliance (%) |
| --- | --- | --- |
| Scrubbing Duration $\geq 2$ minutes | 45 | 90 |
| Coverage of all hand surfaces | 50 | 88 |
| Proper antiseptic usage | 60 | 93 |
| Correct drying technique | 55 | 87 |
| Adherence to aseptic protocol | 70 | 95 |

## Discussion

This audit demonstrates that structured educational interventions can dramatically improve hand-scrubbing compliance in orthopedic surgical settings. Baseline observations revealed common deficiencies in duration, thoroughness, and technique, consistent with findings from other resource-limited healthcare environments [4,5,7].

Live demonstrations proved particularly effective, allowing participants to learn through observation and real-time feedback. Visual reminders, such as posters, reinforced these practices and provided ongoing cues within the scrub area. These results align with prior studies highlighting the efficacy of multimodal hand-hygiene interventions over single approaches [6,8].

Interestingly, while senior staff had higher baseline compliance, the greatest relative improvement was seen in junior staff and OTAs, suggesting that educational interventions are particularly beneficial for less experienced team members. Ensuring that all personnel participate in training is critical for consistent hand hygiene practices.

Enhanced hand hygiene directly contributes to patient safety by reducing the risk of surgical site infections, which is particularly relevant in orthopedic surgery with implantable devices and open fractures [2,3]. The audit demonstrates that low-cost, practical interventions can achieve rapid and substantial improvements.

## Data Availability

All data produced in the present study are available upon reasonable request to the authors

## Limitations

This audit was conducted in a single operating theatre, with a limited follow-up period, preventing assessment of long-term sustainability. Observer presence may have influenced participant behavior (Hawthorne effect).

## Future Recommendations

Regular refresher sessions, continuous visual reminders, and repeated audits should be implemented to sustain high compliance. Expanding the audit across multiple operating theatres or hospitals would provide broader insights into systemic improvements.

## Conclusions

The prospective audit shows that hand-scrubbing compliance in an orthopedic operating theatre can be significantly improved through structured educational interventions, including live demonstrations and visual cues. Such interventions are practical, cost-effective, and directly contribute to patient safety. Routine audits should be incorporated into surgical practice to ensure sustained adherence to WHO hand hygiene standards.

## Appendices

**Table 2:** Hand-Scrubbing Observation Checklist Used for the Prospective Clinical Audit.

| Component | Observation Criteria |
| --- | --- |
| Scrubbing Duration | ≥2 minutes for routine hand wash |
| Coverage | Entire hand surface (palms, dorsum, between fingers, fingertips, thumbs) |
| Forearm Coverage | Up to elbow, no areas missed |
| Antiseptic Use | Correct amount of antiseptic applied |
| Technique | Circular rubbing movements, interlaced fingers, attention to nails |
| Drying | Hands and forearms dried with sterile towel without contamination |
| Glove Donning | Sterile gloves donned immediately after drying |
| Adherence to Asepsis | No contact with non-sterile surfaces before entering OR |

## Acknowledgement

The authors would like to sincerely thank the Head of the Orthopedic Department and the Academic Dean for their guidance, support, and encouragement throughout the conduct of this audit. Their expertise and advice were invaluable in the design, implementation, and review of this study.

## Ethics Statement

Ethical oversight for this study was provided by the Ethical Review Committee of Regional Headquarter Hospital Skardu, Gilgit-Baltistan, Pakistan. The committee reviewed the study protocol and determined that the project constituted a clinical audit of operating theatre practices using anonymized observational data. Therefore, formal ethical approval was waived.

## References

1. World Health Organization. WHO guidelines on hand hygiene in health care. Geneva, Switzerland: WHO Press; 2009.

2. Mangram AJ, Horan TC, Pearson ML, Silver LC, Jarvis WR: Guideline for prevention of surgical site infection, 1999. Hospital Infection Control Practices Advisory Committee. Am J Infect Control. 1999, 27:97–132. 10.1016/S0196-6553(99)70088-2

3. Tanner J, Khan D, Fitzpatrick M, et al.: Hand hygiene compliance in surgery: a systematic review. J Hosp Infect. 2020, 104:1–10. 10.1016/j.jhin.2019.09.024

4. Allegranzi B, Pittet D: Role of hand hygiene in healthcare-associated infection prevention. J Hosp Infect. 2009, 73:305–315. 10.1016/j.jhin.2009.04.019

5. Erasmus V, Daha TJ, Brug H, et al.: Systematic review of studies on compliance with hand hygiene guidelines in hospital care. Infect Control Hosp Epidemiol. 2010, 31:283–294. 10.1086/650451

6. Sax H, Allegranzi B, Chraiti MN, et al.: The World Health Organization hand hygiene observation method. Am J Infect Control. 2009, 37:827–834. 10.1016/j.ajic.2009.07.001

7. Pittet D, Hugonnet S, Harbarth S, et al.: Effectiveness of a hospital-wide programme to improve compliance with hand hygiene. Lancet. 2000, 356:1307–1312. 10.1016/S0140-6736(00)02814-2

8. Luangasanatip N, Hongsuwan M, Limmathurotsakul D, et al.: Comparative efficacy of interventions to promote hand hygiene in hospital: systematic review and network meta-analysis. BMJ. 2015, 351:3728. 10.1136/bmj.h3728

